# Resting Heart Rate predicts all-cause mortality in sub-Saharan African Patients with Heart Failure: Insight from the Douala Heart failure registry (Do-HF)

**DOI:** 10.1101/2020.11.24.20237701

**Authors:** Anastase Dzudie, Blaise Barche, Sidick Mouliom, Ariane Nouko, Fogue Raissa, Jules Njebet, Abang Sarah, Joseph Abah, Armel Djomou, Archange Nzali, Clovis Nkoke, Felicite Kamdem, Samuel Kingue, Karen Sliwa, Andre Pascal Kengne

**Author notes:** **Corresponding authors:** Professor Anastase Dzudie, *Service of Internal Medicine and Cardiology, Douala General Hospital, Cameroon* Po box 4856, Douala, Cameroon.

## Abstract

**Background:** Higher resting heart rate (HR) is associated with mortality amongst Caucasians with heart failure (HF), but its significance has yet to be established in sub-Saharan Africans in whom HF differs in terms of characteristics and etiologies.

**Objectives:** We assessed the association of HR with all-cause mortality in patients with HF in sub-Saharan Africa.

**Methods:** The Douala HF registry (Do–HF) is an ongoing prospective data collection on patients with HF receiving care at four cardiac referral services in Douala, Cameroon. Patients included in this report were followed-up for 12 months from their index admission, for all-cause mortality. We used Cox-regression analysis to study the association of HR with all-cause mortality during follow-up.

**Results:** Of 347 patients included, 343 (98.8%) completed follow-up. The mean age was 64±14 years, 176 (50.7%) were female, and median admission HR was 85 bpm. During a median follow-up of 12 months, 78 (22.7%) patients died. Mortality increased steadily with HR increase and ranged from 12.2% in the lower quartile of HR (≤69 bpm) to 34.1% in the upper quartile of HR (>100 bpm). Hazard ratio of 12-month death per 10 bpm higher heart rate was 1.16 (1.04–1.29), with consistent effects across most subgroups, but a higher effect in participants with hypertension vs those without (interaction p=0.044).

**Conclusion:** Heart rate was independently associated with increased risk of all-cause mortality in this study, particularly among participants with hypertension. The implication of this finding for risk prediction or reduction should be actively investigated.

## Introduction

High resting heart rate (HR) is as a strong predictor for adverse outcomes in several cardiovascular diseases including heart failure (HF) [1–3]. In patients with suspected or proven coronary artery disease, resting HR was shown by Diaz A et al [4] to be a predictor of mortality. In HF Caucasians with reduced or preserved ejection fraction (HFrEF or HFpEF), with or without HF symptoms or signs, high HR has been correlated with adverse outcomes, independently of traditional risk factors [2–4]. Accordingly, clinical trials on limiting HR medications used in HF like beta-blockers and Ivabradine suggest that benefit comes from their effect in reducing HR[5,6]. HR has therefore been considered as a target in HF and limiting HR medications are recommended to be used in HF by the European Society of Cardiology [7]. However, HR may differ by age and HF etiology and possibly other factors, such as ethnicity.

Given HF in sub-Saharan Africans occurs two decades earlier and is predominantly non-ischemic unlike western countries[8], it is uncertain whether HR would be a predictor of adverse outcomes in this specific population. Thus, we carried out this study to evaluate the prognostic value of HR in sub-Saharan African patients with HF. We hypothesized that higher HR would be associated with poor adverse outcomes.

## Methods

### Study design and clinical setting

The Douala HF registry (DoHF) is a prospective, multisite, observational data collection on HF patients that was initiated in 2016 in four cardiology centers in the economic capital of Cameroon, central Africa. Douala is the largest city in Cameroon with a heterogeneous population. The centers were selected on the basis of availability of a clinical cardiologist able to perform echocardiography, and with previous experience in conducting cohort studies. Diagnosis of HF was in accordance with the European society of cardiology (ESC) guidelines [7].

### Inclusion and exclusion criteria

All patients aged 21 years and above with clinical signs and symptoms consistent with congestive HF (i.e., pedal oedema, elevated jugular venous pressure, pulmonary congestion, and tender hepatomegaly) and who were willing to continue follow-up for a minimum of 12 months were included. Written informed consent was obtained from each subject enrolled into the study. Patients were excluded if they refused to give informed consent. Ethical approval was obtained from the Cameroon National Ethics Committee before commencement of the registry by participating institutions, and the study conformed to the principles outlined in the Declaration of Helsinki.

### Data collection

For all patients included in the registry, baseline resting HR was obtained from resting 12-lead ECG tracings undertaken as diagnostic workup. Other baseline characteristics included clinical signs and symptoms (NYHA classification), clinical parameters, transthoracic echocardiography parameters (ejection fraction and information on valvular dysfunction laboratory investigations (serum electrolytes, renal function test), associated cardiovascular risk factors and medications. Follow-up information was obtained from the registry with all-cause mortality being the outcome of interest.

### Statistical analyses

SPSS® version 25 (International business machines corp, New York City) was used for analysis, HR was divided into quartiles based on distribution of baseline HR. Baseline characteristics overall and across HR quartiles are presented means and standard deviation for continuous variables and frequencies and percentages for categorical variables; with group comparisons via one-way analysis of variance and chi square test. Time-to-event analyses were based on cox proportional hazard regression models. The effect of baseline HR on mortality risk was tested in two ways: firstly by computing the hazard ratio and 95% confidence interval for all-cause mortality across increasing quarters of baseline HR, using the lower quarter as reference; secondly, by fitting a continuous predictor to estimate the risk of death associated with a 10 bpm increase in baseline HR, and consistency in the effects across major subgroups assessed via interaction tests. Kaplan Meier estimator was use to illustrate the risk of death during follow-up across quarters of baseline HR, with comparison across quarters based on the logrank test. Statistical significance was considered for p < 0.05. Missing data as well as participants who were considered loss to follow up were excluded from Cox proportional regression model analysis

## Results

### Patient characteristics

Of the 347 patients included at baseline, 343 (98.8%) had complete follow-up data. Baseline characteristics of these patients by HR quarters are presented in *Table 1*. Overall, the mean age was 64±14 years. Likewise, patients with the highest resting HR were younger and were more likely to have a history of HF in the past two years compared to those with lower resting HRs. Patients with higher resting HR were also likely to present with lower LVEF, lower estimated glomerular filtration rate when compared to the lower heart groups. There was no difference across the groups with regards to use of beta blockers, ACE inhibitors and angiotensin receptor blockers or loop diuretics

**Table 1.** Demographics, Medical History, clinical and paraclinical findings, and Medication Use by Heart Rate Group

| parameters | HR ≤69 bpm<br>(n=90) | HR 70 -85bpm<br>(n=94) | HR 86 -<br>100bpm (n=81) | HR >100 bpm<br>(n=82) | Total (n = 347) | p-value* |
| --- | --- | --- | --- | --- | --- | --- |
| Age(years) | 64.81(13.49) | 64.14(14.78) | 66.04(12.22) | 61.28(15.11) | 64.08(14.02) | 0.19 |
| Sex(male) | 45(50%) | 51(54.3%) | 39(48.1%) | 37(45.1%) | 172(49.6%) | 0.39 |
| Prior Hx of HF | 20(22.2%) | 26(27.7%) | 28(34.6%) | 28(27.5%) | 102(29.4%) | 0.051 |
| Hypertension | 61(67.8%) | 59(62.8%) | 60(74.4%) | 46(56.1%) | 226(65.1%) | 0.32 |
| Diabetes mellitus | 6(6.7%) | 8(8.5%) | 14(17.3%) | 15(18.3%) | 43(12.1%) | 0.006 |
| Dyslipidemia | 14(15.6%) | 12(12.8%) | 16(19.8%) | 3(3.7%) | 45(13.0%) | 0.12 |
| Inpatient | 6(6.7%) | 20(21.3%) | 19(23.5%) | 38(46.3%) | 83(23.9%) | <0.001 |
| Out patient | 84(93.3%) | 74(78.7%) | 62(76.5%) | 44(53.7%) | 264(76.1%) | 0.004 |
| Heart rate | 61.15(6.81) | 77.94(4.60) | 92.42(4.39) | 120.95(16.47) | 87.13(23.70) | <0.001 |
| SBP (mm Hg) | 133.0(29.71) | 134.85(28.17) | 136.20(31.89) | 129.18(32.62) | 133.35(30.51) | 0.49 |
| BMI(Kg/m<sup>2</sup>) | 27.02(6.01) | 26.59(5.07) | 26.64(5.67) | 27.03(6.92) | 26.82(5.91) | 0.98 |
| DBP (mm Hg) | 77.44(17.06) | 82.03(17.31) | 85.90(20.68) | 85.01(18.25) | 82.45(18.52) | 0.003 |
| NYHA I and II | 52(57.8%) | 55(58.5%) | 25(30.9%) | 31(37.8%) | 163(47.0%) | 0.001 |
| NYHA III and IV | 38(42.2%) | 39(41.5%)) | 56(69.1%) | 51(62.2%) | 184(53.0%) | 0.16 |
| LVEF (%) | 46.17(16.71) | 45.06(15.33) | 38.07(17.06) | 35.93(15.26) | 41.56(16.61) | <0.001 |
| HFrEF | 26(28.9%) | 37(39.4%) | 48(59.3%) | 51(62.2%) | 162(46.7%) | <0.001 |
| HFmrEF | 27(30%) | 20(21.3%) | 14(17.3%) | 19(23.2%) | 80(23.1%) |  |
| HFpEF | 37(41.1%) | 37(39.4%) | 26(23.5%) | 12(14.6%) | 105(30.3%) |  |
| Creatinine, mg/dl | 1.31(0.56) | 1.49(0.79) | 1.44(0.98) | 1.61(1.07) | 1.47(0.89) | 0.10 |
| Sodium, mmol/l | 138.35(6.26) | 138.12(8.08) | 137.47(5.38) | 136.21(5.75) | 137.53(6.51) | 0.074 |
| Potassium, mmol/l | 4.21(0.66) | 4.25(0.69) | 4.10(0.71) | 4.07(0.65) | 4.16(0.68) | 0.16 |
| eGFR ml/min/1.73m<sup>2</sup> | 65.45(242.3) | 61.27(23.87) | 63..38(25.28) | 58.44(27.07) | 61.98(25.17) | 0.20 |
| Hemoglobin(mg/dl) | 11.83(1.86) | 11.39(2.06) | 11.29(2.02) | 11.84(2.12) | 11.59(2.03) | 0.95 |
| ACEi (yes) | 58(64.4%) | 64(68.1%) | 56(69.1%) | 53(65.4%) | 231(66.8%) | 0.85 |
| ARB (yes) | 21(23.3%) | 16(17.0%) | 15(18.5%) | 14(17.3%) | 66(19.1%) | 0.38 |
| MRA(yes) | 35(38.9%) | 43(45.7%) | 41(50.6%) | 36(44.4%) | 155(44.8%) | 0.36 |
| Beta blockers(yes) | 57(63.3%)) | 69(73.4%) | 56(69.1%) | 56(69.1%) | 238(68.8%) | 0.55 |
| CCB (yes) | 18(20.0%) | 8(8.5%) | 5(6.2%) | 5(6.2%) | 36(10.4%) | 0.003 |
| Diuretics(yes) | 83(93.3%) | 84(89.4%) | 76(93.8%) | 79(97.5%) | 322(933.3%) | 0.16 |
| Atrial fibrillation/flutter | 15(17.9%) | 19(21.3%) | 19(24.4%) | 42(52.5%) | 95(28.7%) | <0.001 |

\*p-value for linear trend
Presented are mean (SD) and median (25<sup>th</sup>, 75<sup>th</sup> percentiles) for continuous variables and n (%) for categorical variables. Statistics are computed from non-missing values, the number of which may vary from variable to variable.
ACE = angiotensin-converting enzyme; ARB = angiotensin receptor blocker, CCB = calcium channel blockers, eGFR = estimated glomerular filtration rate, ECG = elcetrocardiogram, LVEF = left ventricular ejection fraction, HFmrEF= heart failure with moderately reduced ejection fraction, HFpEF= heart failure with preserved ejection fraction, HFrEF = heart failure with reduced ejection fraction, HR = hear rate, Hx of HF = history of heart failure

### Heart Rate and Survival

During a mean follow-up of 12 months, 78 patients died. All-cause mortality at 1 year was significantly lower in the group with HR of <69 beats/min (9%) when compared to patients with higher HR (p =0.001) (Figure 1). The pattern was similar in participant in sinus rhythm at baseline (logrank p=0.005) but only borderline in participants with atrial fibrillation or flutter at baseline (logrank p=0.089).

**Figure 1:**
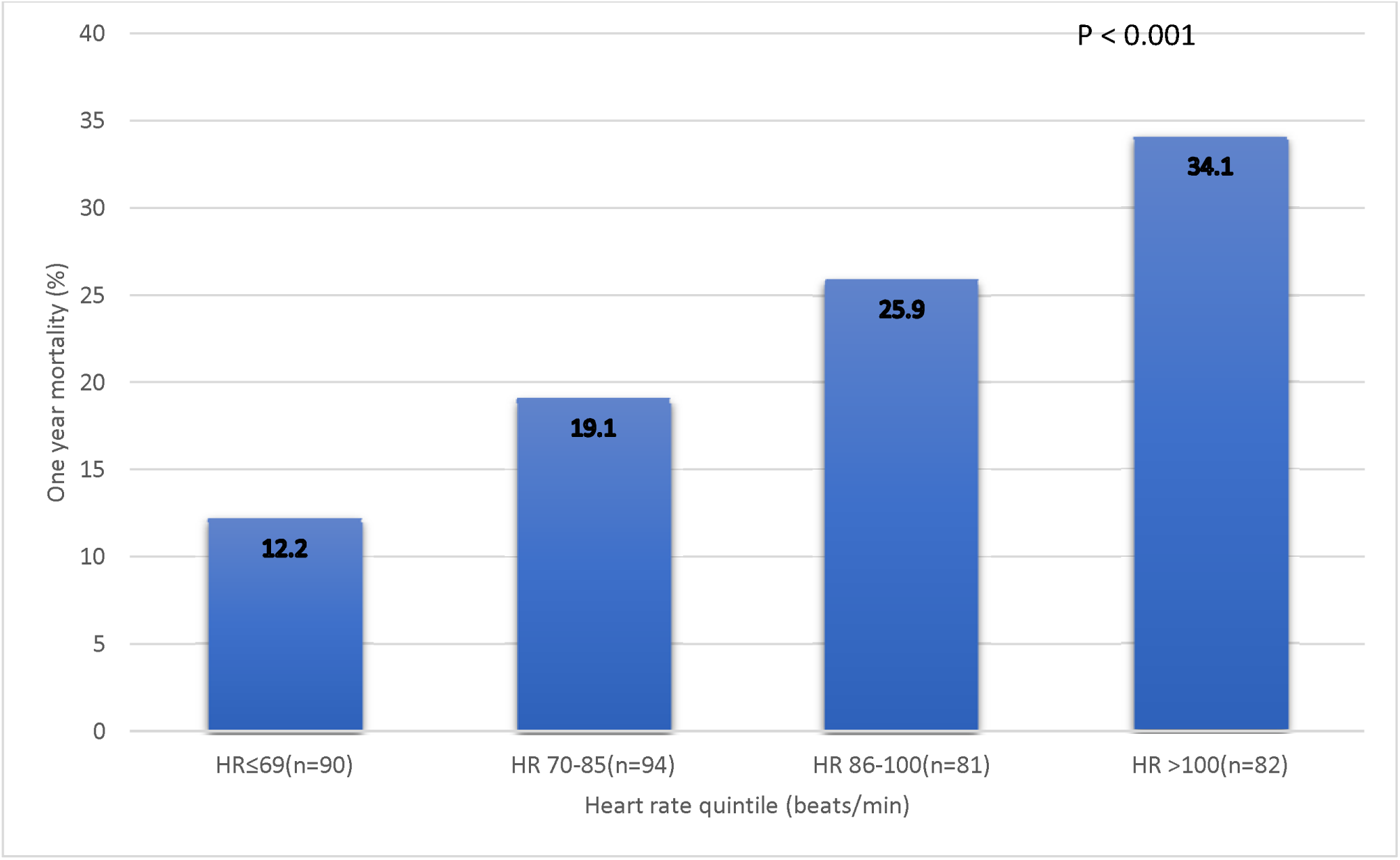
One-year all-Cause Mortality by baseline Heart Rate in patients with heart failure (the histogram)

Age-adjusted survival for the different HR groups is displayed in Fig 2, showing that those with higher HRs had shorter survival when compared to participants in the lower HR quarter. This pattern was observed in the overall sample (logrank p<0.001), and within subgroups defined by the baseline presence or absence of atrial fibrillation/flutter, and presence or absence of sinus rhythm. However, due to small numbers, differences across HR quarters within subgroups were not always significant. Survival curves for participants with atrial fibrillation/flutter, and those in sinus rhythm at baseline are shown in Figure 3. After adjusting for age, hypertension, diabetes mellitus, left ventricular ejection fraction, renal function cardiac rhythm, beta blocker use, and congestion; and using the lower quarter as reference in the overall sample, the hazard ratio of all-cause mortality was 2.45 (95%CI: 0.93-6.44) in the second quarter, 1,70 (0.63-4.65) in the third quarter and 2.98 (1.13-7.89) in the top quarter. Small numbers precluded similar analyses in subgroups. With similar level of adjustment for confounders, the hazard ratios for every 10-beat increase in HR in the overall sample and across subgroups are represented in Figure 4. In the overall sample, each 10-beat higher heart rate was associated with a 16% (95%CI 4-29%) higher risk of 12-month mortality. In addition, congestion at baseline was associated with a 247% (95-518%) higher risk of mortality, while age, ejection fraction, hypertension, diabetes mellitus, use of betablockers and rhythm were not associated with risk of death during follow-up. Across complementary subgroups defined by gender (men vs. women), age (<60 vs ≥60 years), setting of initial follow-up (inpatient vs outpatients) renal function (altered vs unaltered), rhythm (sinus vs. atrial flutter or fibrillation), ejection fraction (reduced, mid-range, preserved), use of betablocker (yes vs no), congestion (yes vs. no), each 10 unit higher baseline heart rate was associated with similar increase in the risk of 12-month mortality, with no evidence of statistical interaction (all interaction p≥0.31); Figure 4. There was however a significant interaction by status for hypertension on the effect of heart rate on all-cause mortality risk, with indication that the effect was more pronounced in participants with a history of hypertension [hazard ratio 1.30 (95%CI: 1.16-1.45)] than in those without [1.08 (0.96-1.23)], interaction p=0.044.

**Figure 2:**
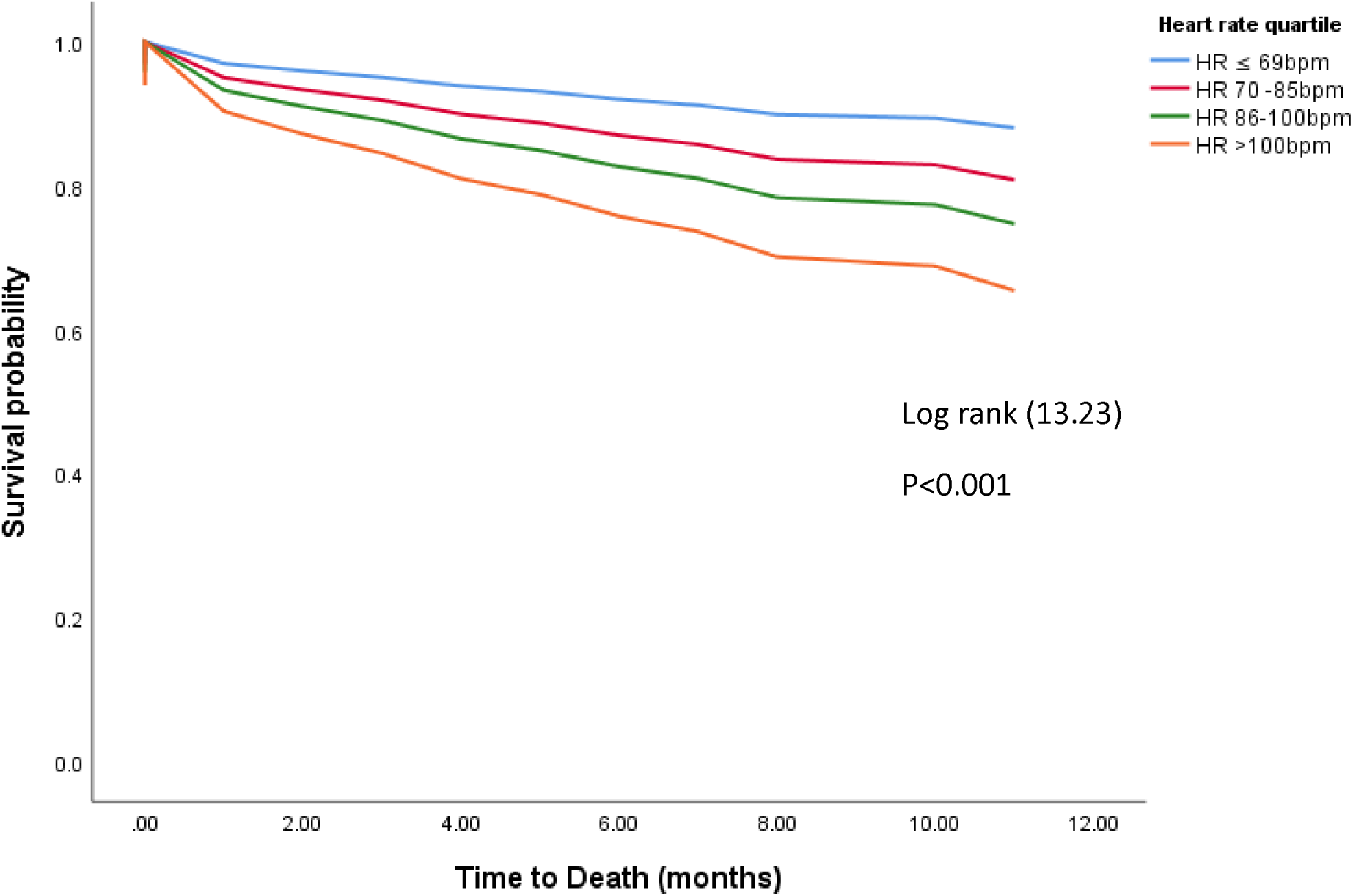
Kaplan-Meier survival curves showing that all-cause mortality increases with increasing resting heart rate.

**Figure 3a:**
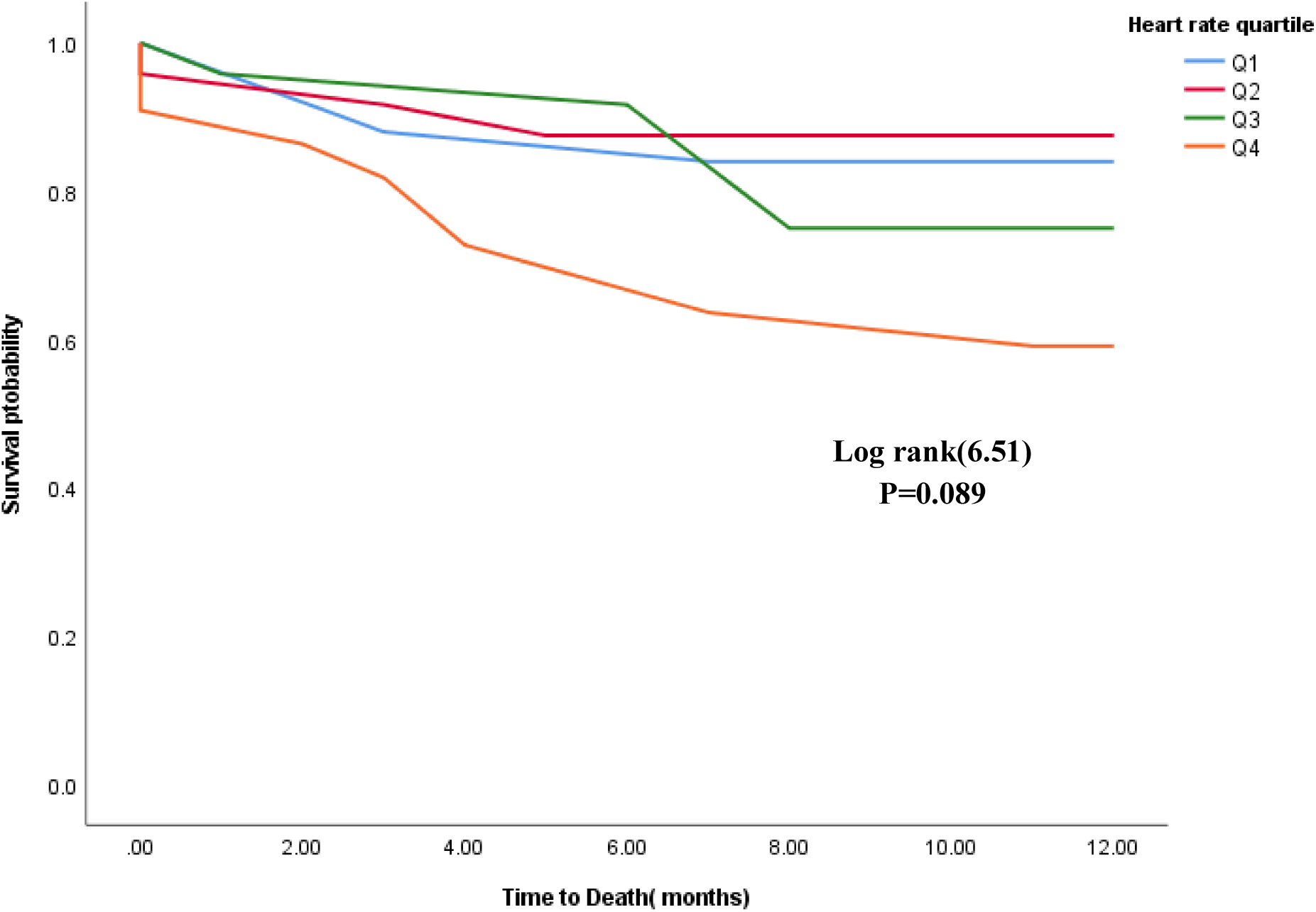
Survival curve in subgroup of patients in atrial fibrillation/flutter

**Figure 3b:**
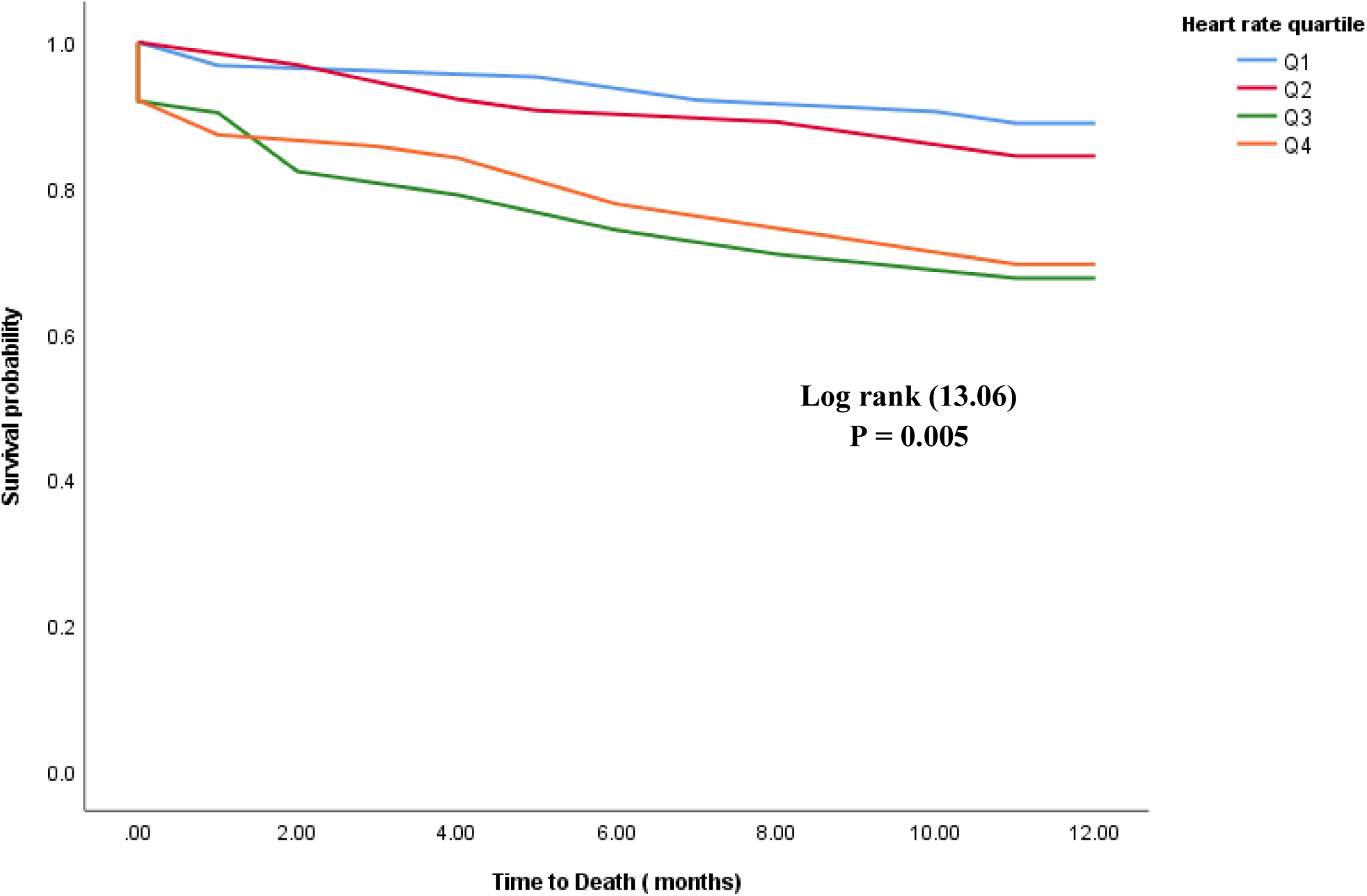
Survival curve in subgroup of patients in in sinus rhythm

**Figure 4:**
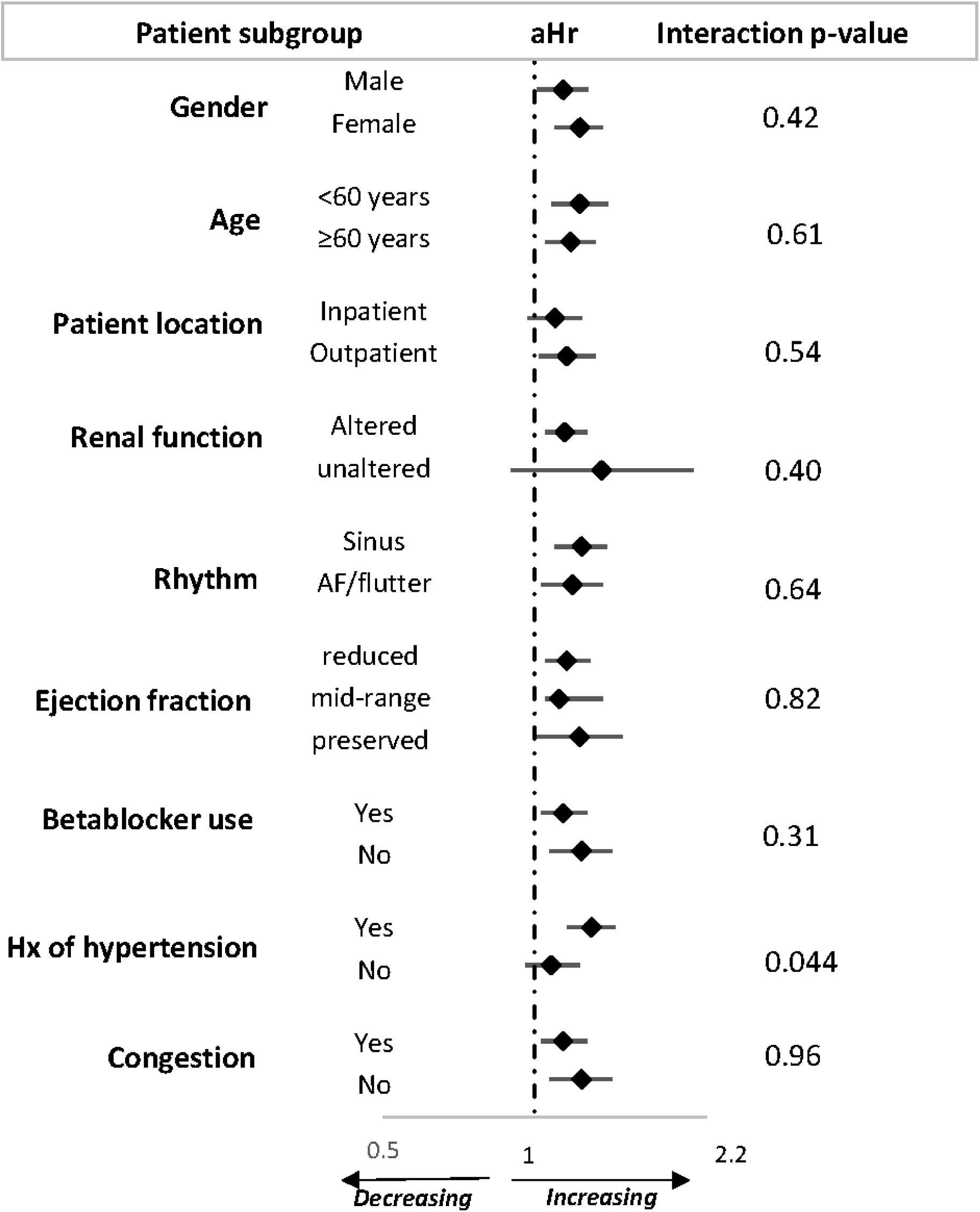
Adjusted hazard ratio and 95% confidence interval for each 10 unit higher heart rate in relation with all-cause mortality, across major subgroups of participants AF = atrial fibrillation, aHr = adjusted hazards ratio/ 10 beat increase in heart ratre

## DISCUSSION

This study reveals that sub Saharan African HF patients with higher resting HR have a significantly higher risk of mortality when compared to patients with lower resting HR, with each 10-unit higher heart rate conferring a 16% higher risk of 12-month mortality. While being mostly consistent across most subgroups of participants, this effect appeared to be more important in participants with hypertension at baseline, compared to those without, resulting in significant statistical interaction. To our knowledge this is one of the rare studies on the predictive significance of HR rate for all-cause mortality in a cohort of HF patients in sub-Saharan Africa.

Our results are in agreement with those reported in several studies [9–12] most importantly these results are in line with two previous post-hoc analysis from the THESUS-HF study in which we evaluated the predictive value of several patient factors including ECG abnormalities on 6-month all-cause mortality in patients with heart failure[13,14]. Although the THESUS-HF did not focus on HR as a principal predictor of mortality, it was purely an African study [8], with patients of similar profile to those in the current study. Notably, the effect of increased HR was marked in the subgroup of patients with hypertension. Despite the variations of etiologies and clinical characteristics of HF globally, a similar effect has been reported in several studies[15–17]. It shall be noted that hypertension has been reported as one of the common causes of heart failure in SSA [8]. The enhanced effect of HR on mortality risk in the presence of hypertension can be explained by the elevated risk of arterial stiffness associated with increased HR and hypertension[18,19], which in turn has been reported as a significant predictor of mortality [20].

Increased resting HR is a regular finding in HF. This results from a sustained vagal nerve activity inhibition and a hypersympathetic activity [21,22]. As HF progresses, there is an enhancement of the hypersympathetic activity representing an attempt to overcome the hemodynamic failure due to HF and to restore normal cardiac output; but this has been essentially studied in Caucasians [23–25]. Regional differences in the patient characteristics, management, and outcomes of patients with HF have been described [26,27]. Also, differences in cardiovascular drug responses by different ethnic groups should be considered [28]. The ESC recommends decreasing the HR in HF by adding ivabradine if beta-blockers do not attain the optimal HR [7]. That only 68.8% of our patients received beta-blockers and median HR remains high is an indicator of non-optimal therapy. Similarly these results strengthens the fact that despite variations in distribution of cardiovascular risk factors and etiologies of HF across the globe [27,29], decreasing the HR to optimal values will as well improve outcome in HF patients in sub Saharan Africa. Our study has some weaknesses. First, we used single measurements of HR, assuming that they were representative of the usual HR of the patient. Actually, it is possible that resting HR might not be the best surrogate of HR during daily activities, a more accurate assessment would be through prolonged ambulatory ECG monitoring. However, Johansen et al [32] described a good correlation between resting HR and mean rates on 24-hour monitoring in the general population as well as in ambulatory patients with HF [33]. Secondly, our study group consisted of a general population seen in cardiology units, who may not necessarily represent the general population of patients with HF that are sometimes managed by internists and general practitioners. To our knowledge, this is the first report on the prognostic impact of HR in HF in sub Saharan African black patients, a relatively young population when compared with Caucasians and with predominantly non-ischemic etiology of HF. The fact that HR was obtained from resting 12-lead ECG tracings undertaken as diagnostic workup is a very important strength of our study. Indeed, in several other HF registries on the prognostic impact of HR in HF, previous exact timing of HR measurement was unclear.

## Conclusions

In African patients with HF, resting HR is an independent risk factor for death, with a significantly higher effect in the subgroup of patients with hypertension, in line with reports from other regions around the world. As described by previous studies, the prescription of available therapies with proven efficacy in reducing deaths was rather suboptimal, especially for beta blockers. While additional studies are still warranted to determine if simple interventions designed to lower the HR will improve outcome in this population, optimization of HF therapy with prescription of beta blockers is urgent.

## Data Availability

The data that support the findings of this study are available on request from the corresponding author.
The data are not publicly available due to privacy or ethical restrictions.

## Acknowledgements

The authors wish to thank all the doctors, nurses and patients who participated in the registry.

## Authors’ contributions

I. Conception and design: Dzudie Anastase, Andre Pascal Kengne, Blaise Barche
II. Administrative support: Dzudie Anastase
III. Provision of study materials or patients: Anastase Dzudie, Sidick Molioum, Felicite Kamdem, Jules Njebet, Abang Sarah, Joseph Abah, Armel Djomou, Archange Nzali
IV. Collection and assembly of data: Fogue Raissa, Ariane Nouko
V. Data analysis and interpretation: Dzudie Anastase, Andre Pascal Kengne, Blaise Barche, Fogue Raissa
VI. Manuscript writing: All authors.
VII. Final approval of manuscript: All authors.

## Ethical statement

The authors are accountable for all aspects of the work in ensuring that questions related to the accuracy or integrity of any part of the work are appropriately investigated and resolved

## Data sharing statement

The data that support the findings of this study are available on request from the corresponding author. The data are not publicly available due to privacy or ethical restrictions.

